# High Impedance Microstrip Resonators and Transceiver Arrays for Ultrahigh Field 10.5T MR Imaging

**DOI:** 10.1101/2025.01.02.25319876

**Authors:** Xiaoliang Zhang, Komlan Payne, Yunkun Zhao

## Abstract

Multichannel transceiver coil arrays are needed to enable parallel imaging and B1 manipulation in ultrahigh field MR imaging and spectroscopy. However, the design of such transceiver coils and coil arrays often faces technical challenges in achieving the required high operating frequency at the ultrahigh fields and sufficient electromagnetic (EM) decoupling between resonant elements. In this work, we propose a high impedance microstrip transmission line resonator (HIMTL) technique that has unique high frequency capability and adequate EM decoupling without the use of dedicated decoupling circuits. To validate the proposed technique for the ultrahigh field 10.5T applications, a two-channel high impedance microstrip array with the element dimension of 8cm by 8cm was built and tuned to 447 MHz, Larmor frequency of proton at 10.5T, for signal excitation and reception. Bench tests and numerical simulations were performed to evaluate its feasibility and performance. The results show that the proposed high impedance microstrip technique can be a simple and robust way to design high frequency transceiver coils and coil arrays for ultrahigh field MR applications.

## Introduction

Whole-body MR imaging at ultrahigh magnetic fields, such as 7T and 10.5T, offers significant advantages, including improved signal-to-noise ratio (SNR), which is crucial for achieving high spatial and temporal resolutions as well as enhanced spectral dispersion [1-9]. However, as the magnetic field strength increases, the corresponding Larmor frequency rises proportionally, posing considerable challenges in designing highly efficient RF coils—key components for MR signal excitation and reception. These challenges become even more pronounced in human imaging due to the requirement for large RF coils [10-25].

At such high frequencies, for example, 447 MHz at 10.5T, tuning RF coils is particularly difficult due to their large inductance and non-negligible parasitic capacitance [26]. Furthermore, the increased radiative behavior of resonant circuits at high frequencies significantly degrades the quality factor (Q-factor) of RF coils and exacerbates electromagnetic (EM) coupling between resonant elements in coil arrays, making multichannel coil array design particularly challenging [27-34].

Microstrip resonators, known for their high-frequency capability, high Q-factors, and reduced radiation losses, have been widely utilized in high-field and ultrahigh-field MR applications [35-42]. Additionally, it has been shown that high-impedance coil designs can reduce EM coupling between array elements, as demonstrated in 1.5T NMR phased arrays [43, 44]. Recent studies have further validated this approach for ultrahigh-field applications, including 7T [45-50].

In this work, we propose a novel microstrip transmission line resonator design that incorporates a unique circuit configuration to achieve both high resonant frequency and high impedance. This design combines the high-frequency capability of microstrip coils with the excellent EM decoupling performance of high-impedance coils, addressing the challenges associated with multichannel coil arrays at the ultrahigh field 10.5T. The proposed technique was validated through numerical simulations and standard RF measurements on a prototype 2-channel high-impedance microstrip array.

The results demonstrate that the proposed high-impedance microstrip coil technique is a practical and efficient solution for designing multichannel coil arrays at ultrahigh magnetic fields, specifically for 10.5T MR imaging.

## Methods

Two high impedance microstrip resonators with a dimension of 8cm by 8 cm were designed and constructed. The 3mm copper tape was used for making the strip conductor. The substrate material is Teflon, and its thickness is ∼2mm. The coil schematics is shown in **Fig 1**. Ct and Cm are tuning and matching capacitors, respectively. This coil has two resonant modes. The high frequency modes possess high impedance feature and thus decoupling advantages, and therefore, is used for this 10.5T proton imaging application. The low frequency mode may not have a high impedance, and stronger coupling between resonant elements should be expected. **Fig 2** shows the photograph of the prototype high impedance microstrip coils set up in a plane and around a human head phantom. The coil’s resonant frequency, Q-factors, decoupling performance were tested on a network analyzer through the measurement of scattering parameters. The coil array was further investigated using numerical simulation tool for its B1 distribution and efficiency of each channel and its decoupling performance.

**Figure 1.**
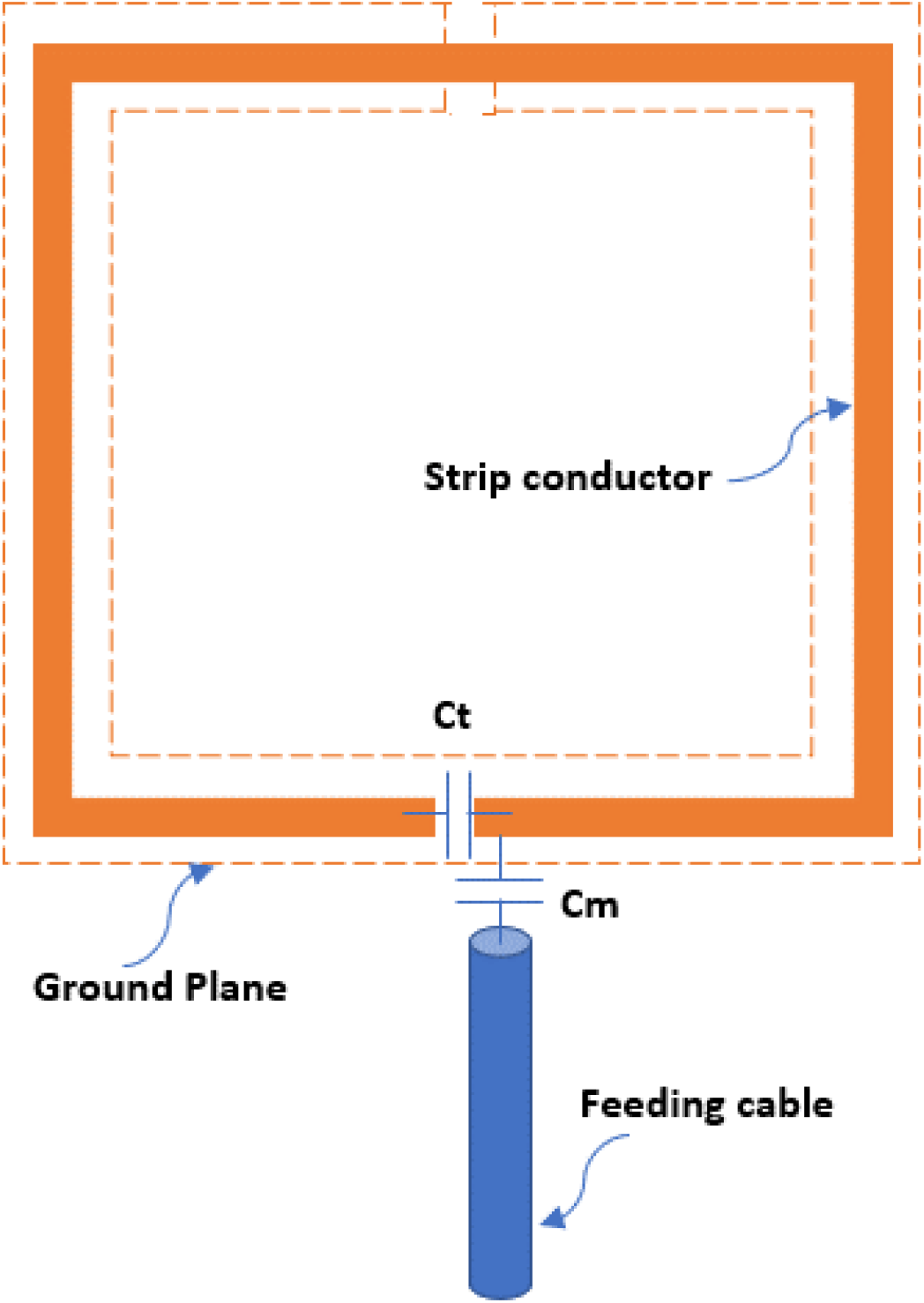
The schematics of the proposed high impedance microstrip RF coil operating for 10.5T proton imaging applications. Ct and Cm are tuning and matching capacitors, respectively. The coil size is 8cm by 8cm. Width of the strip conductor is 3mm while thickness of the Teflon substrate is 2mm. The coil can be easily tuned to the high frequency of 447MHz.

**Figure 2.**
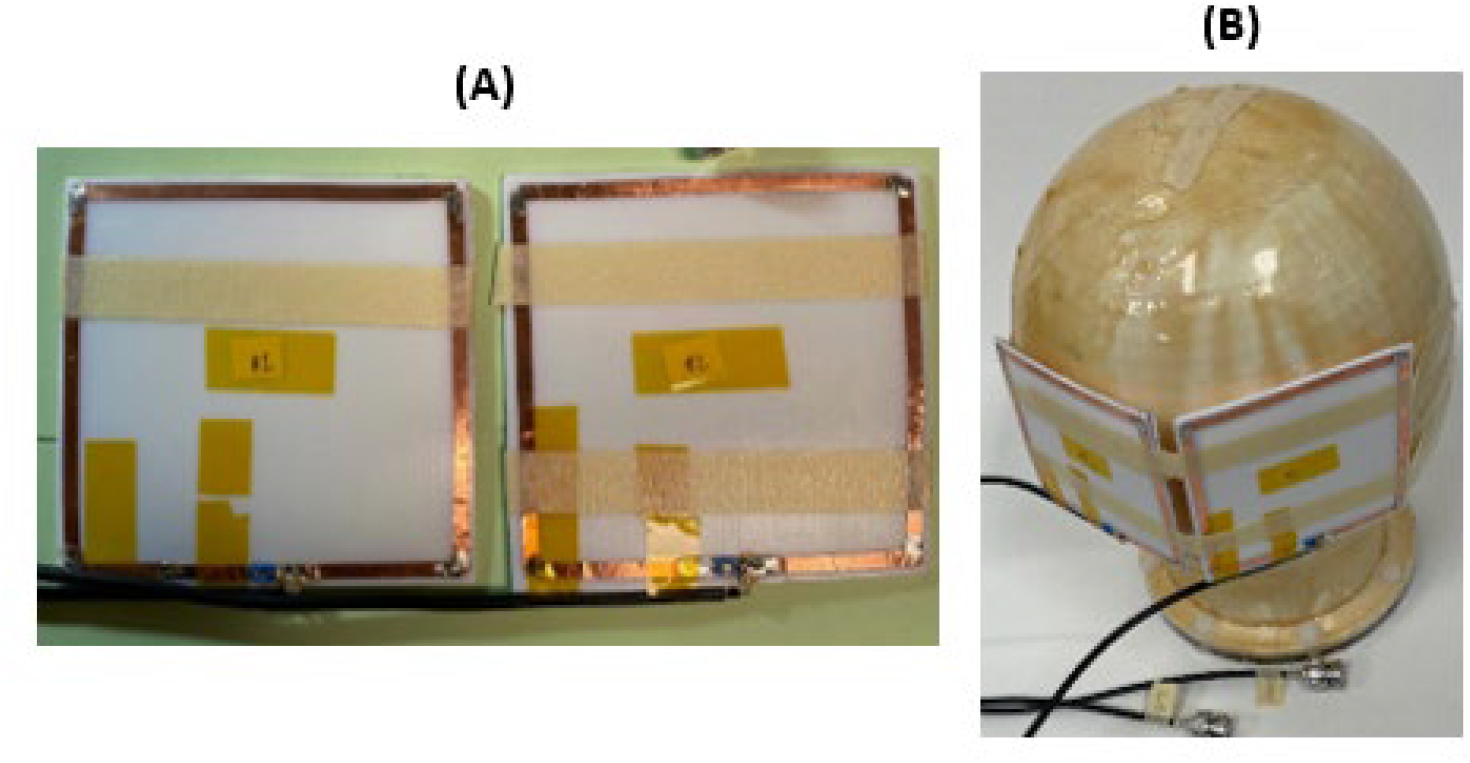
The photograph of the prototype high impedance microstrip coils set up in a plane (**A**) and around a human head phantom (**B**)

## Results

The high impedance microstrip was conveniently tuned to 447MHz level and its input impedance was matched to the system’s 50 ohm with a tunable capacitor. The decoupling performance was tested in two different gaps between the two resonant elements, 10 mm and 2mm. **Fig 3** shows the results of S11 and S21 measurement of the 10.5T high impedance microstrip array when it’s loaded with the human head phantom. The high frequency mode (447MHz) demonstrates robust decoupling of -21 dB between two channels when the two elements are 10mm apart, while the low frequency mode (at 116MHz) shows stronger coupling with a S21 of ∼ -7.6 dB. Testing with a smaller gap of 2mm shows the low frequency had an unacceptable decoupling, making its resonant peak split, as shown in **Fig 4**. Therefore, unlike the high frequency mode, the low frequency mode does not have the self-decoupling capability. A numerical model of the proposed high impedance microstrip coil was built using Ansys simulation software. In the numerical modeling and simulation as shown in **Fig 5**, well defined B1 distribution of each channel was observed, indicating the excellent decoupling performance of the high frequency mode. The calculated B1+ value, with 2uT/sqrt W maximum, of the coil elements is also in a reasonable range.

**Figure 3.**
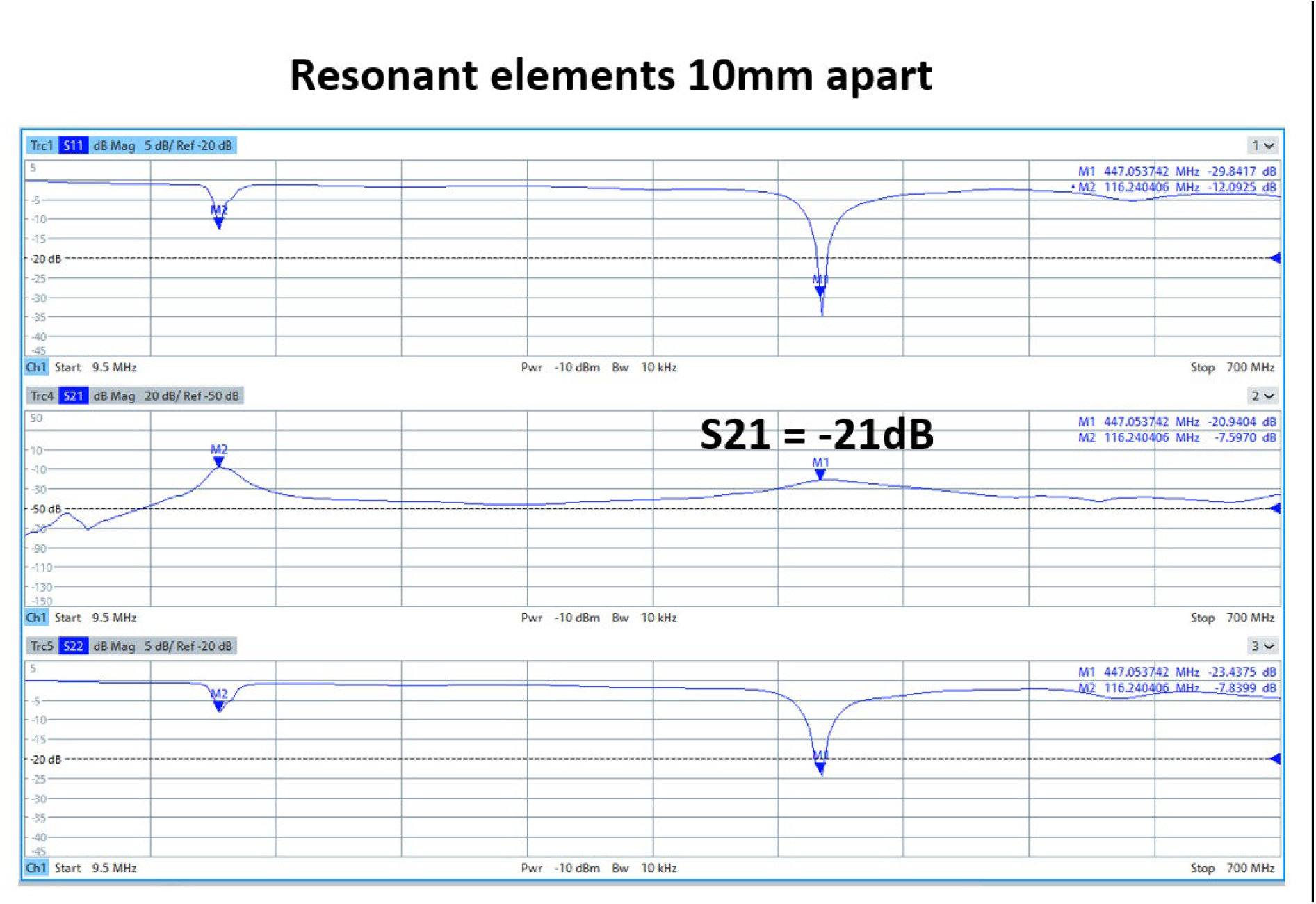
S-parameter measurement of the proposed high impedance microstrip array at 447MHz. The results demonstrate the excellent decoupling performance of the high frequency mode (at 447MHz) while the low frequency mode is weakly coupled. The gap between the coil element is 10mm.

**Figure 4.**
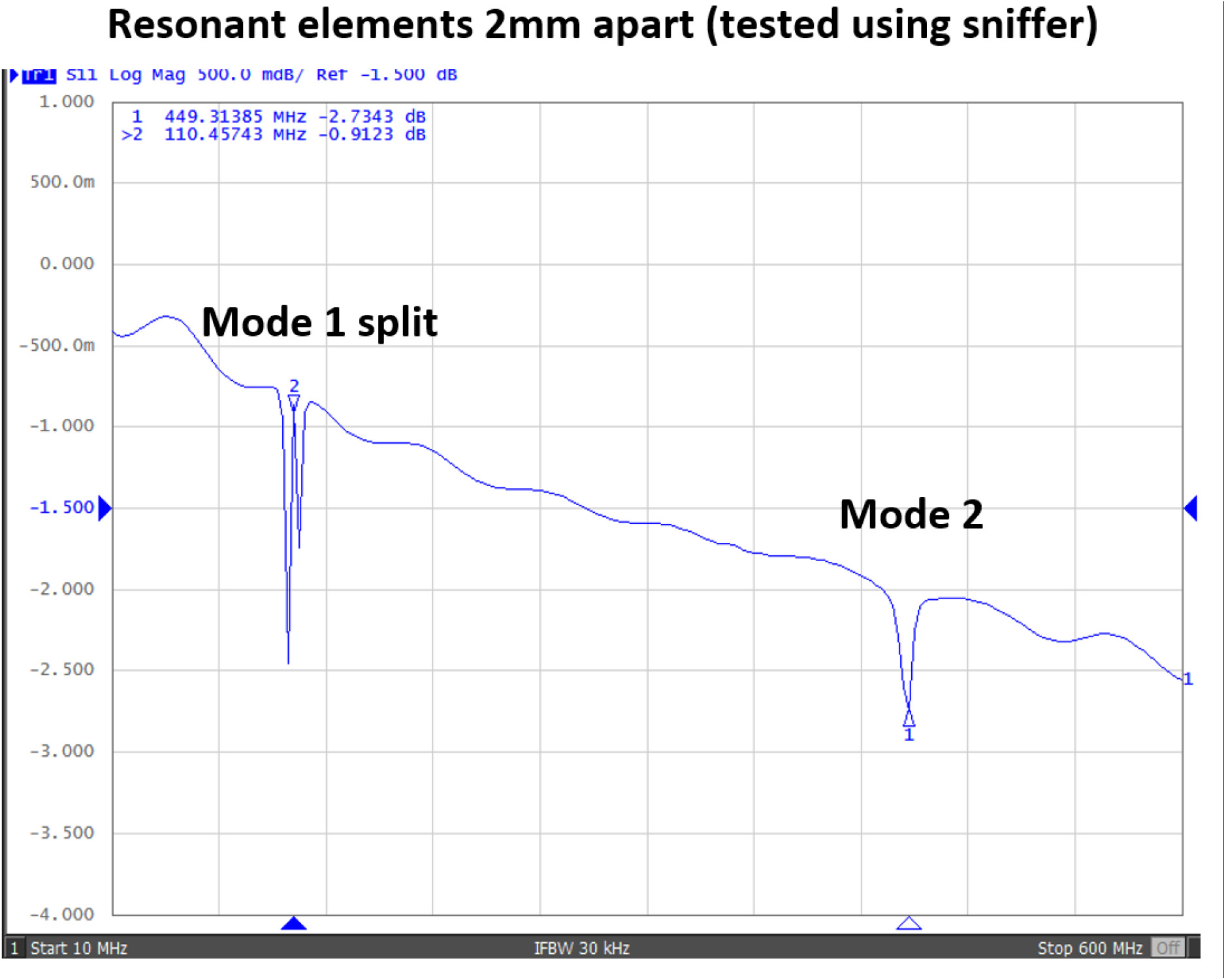
S-parameter measurement of the proposed high impedance microstrip array when the resonant elements are placed 2mm apart. The split peak of mode 1 is observed due to strong coupling, while the high frequency mode still maintains decoupled.

**Figure 5.**
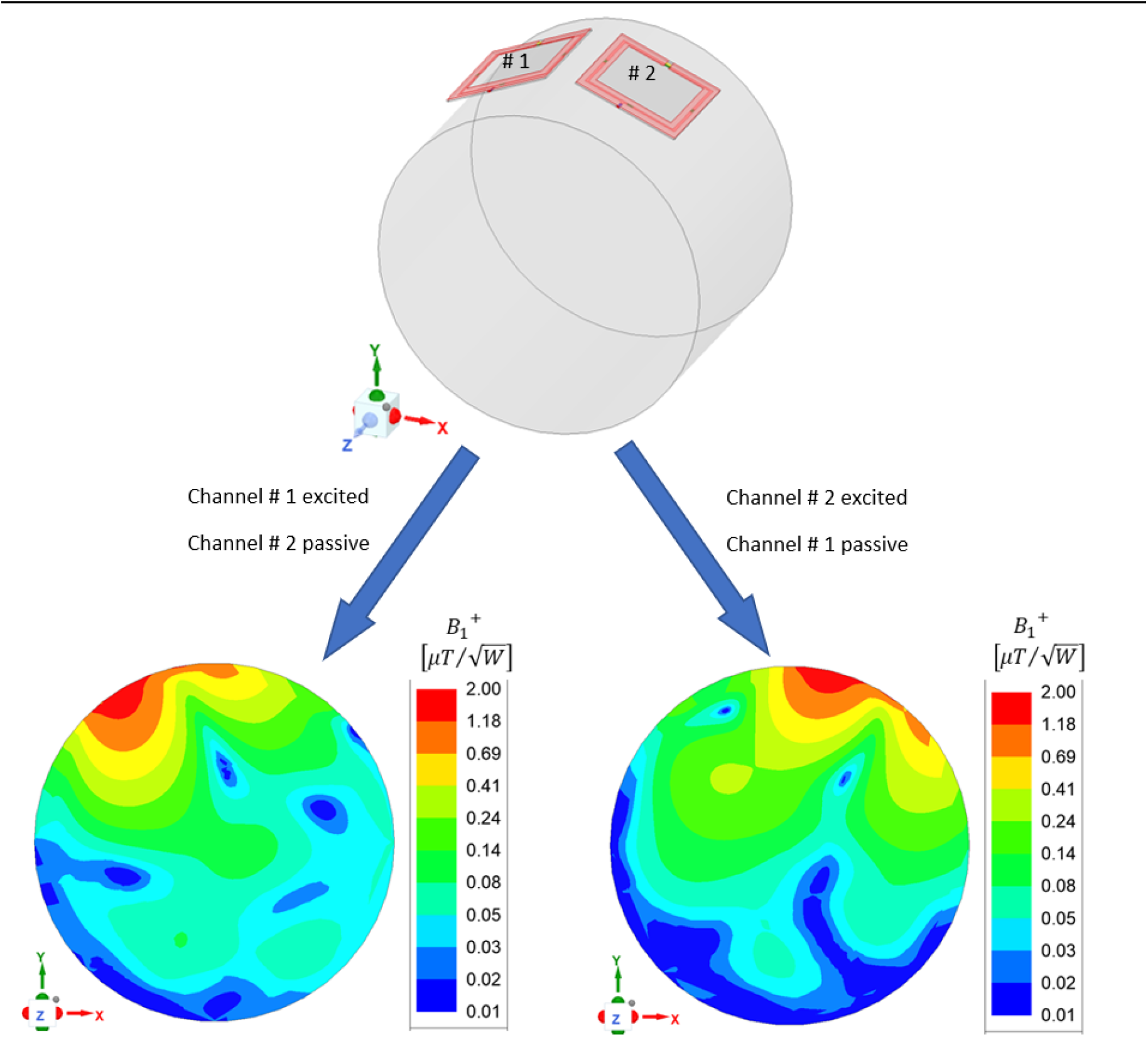
Numerical modeling and simulation of the proposed high impedance microstrip array for 10.5T MR imaging. Well-defined B1 distribution of each channel indicates the excellent decoupling performance of the high frequency mode. The calculated B1+ of the high impedance microstrip coil is also in a reasonable range.

## Conclusion

A high impedance microstrip coil array was designed and tested for 10.5T MR applications. The coil design technique demonstrates its excellence in high frequency operation and element decoupling performance at 447MHz. This technique provides a simple and robust way to design high frequency transceiver arrays for ultrahigh field MR applications.

## Data Availability

All data produced in the present work are contained in the manuscript

## Acknowledgements

This work is supported in part by the NIH under a BRP grant U01 EB023829 and by the State University of New York (SUNY) under SUNY Empire Innovation Professorship Award.

